# Clinician-Led Code-Free Deep Learning for Detecting Papilloedema and Pseudopapilloedema Using Optic Disc Imaging

**DOI:** 10.1101/2025.06.25.25330265

**Authors:** Riddhi Shenoy, Gurtek Singh Samra, Rishi Sekhri, Ha-Jun Yoon, Seema Teli, Ian DeSilva, Zhanhan Tu, Gail DE Maconachie, Mervyn G Thomas

## Abstract

**Importance:** Differentiating pseudopapilloedema from papilloedema is challenging, but critical for prompt diagnosis and to avoid unnecessary invasive procedures. Following diagnosis of papilloedema, objectively grading severity is important for determining urgency of management and therapeutic response. Automated machine learning (AutoML) has emerged as a promising tool for diagnosis in medical imaging and may provide accessible opportunities for consistent and accurate diagnosis and severity grading of papilloedema.

**Objective:** This study evaluates the feasibility of AutoML models for distinguishing the presence and severity of papilloedema using near infrared reflectance images (NIR) obtained from standard optical coherence tomography (OCT), comparing the performance of different AutoML platforms.

**Design, setting and participants:** A retrospective cohort study was conducted using data from University Hospitals of Leicester, NHS Trust. The study involved 289 adults and children patients (813 images) who underwent optic nerve head-centred OCT imaging between 2021 and 2024. The dataset included patients with normal optic discs (69 patients, 185 images), papilloedema (135 patients, 372 images), and optic disc drusen (ODD) (85 patients, 256 images). AutoML platforms - Amazon Rekognition, Medic Mind (MM) and Google Vertex were evaluated for their ability to classify and grade papilloedema severity.

**Main outcomes and measures:** Two classification tasks were performed: (1) distinguishing papilloedema from normal discs and ODD; (2) grading papilloedema severity (mild/moderate vs. severe). Model performance was evaluated using area under the curve (AUC), precision, recall, F1 score, and confusion matrices for all six models.

**Results:** Amazon Rekognition outperformed the other platforms, achieving the highest AUC (0.90) and F1 score (0.81) in distinguishing papilloedema from normal/ODD. For papilloedema severity grading, Amazon Rekognition also performed best, with an AUC of 0.90 and F1 score of 0.79. Google Vertex and Medic Mind demonstrated good performance but had slightly lower accuracy and higher misclassification rates.

**Conclusions and relevance:** This evaluation of three widely available AutoML platforms using NIR images obtained from standard OCT shows promise in distinguishing and grading papilloedema. These models provide an accessible, scalable solution for clinical teams without coding expertise to feasibly develop intelligent diagnostic systems to recognise and characterise papilloedema. Further external validation and prospective testing is needed to confirm their clinical utility and applicability in diverse settings.

**Key Points:** **Question:** Can clinician-led, code-free deep learning models using automated machine learning (AutoML) accurately differentiate papilloedema from pseudopapilloedema using optic disc imaging?

**Findings:** Three widely available AutoML platforms were used to develop models that successfully distinguish the presence and severity of papilloedema on optic disc imaging, with Amazon Rekognition demonstrating the highest performance.

**Meaning:** AutoML may assist clinical teams, even those with limited coding expertise, in diagnosing papilloedema, potentially reducing the need for invasive investigations.

## Introduction

Papilloedema, the swelling of the optic disc due to raised intracranial pressure (ICP), is a critical finding in the diagnosis of various neurological conditions, including idiopathic intracranial hypertension, brain tumours, and venous sinus thrombosis. Early detection is essential to prevent associated morbidity or mortality and to guide appropriate medical intervention (1, 2). However, distinguishing papilloedema from pseudopapilloedema, which can be caused by conditions such as optic disc drusen (ODD), is clinically challenging (3). Accurate diagnosis is crucial to avoid unnecessary invasive procedures, such as lumbar punctures or neuroimaging, which carry risks and financial costs, and to ensure appropriate management for patients.

After establishing a diagnosis of papilloedema, its severity is documented using the Frisén grading system, which is used alongside patient symptomatology and neuroimaging to dictate the urgency of intervention and subsequent therapeutic response. Frisen grading is performed using retinal photographs or on ophthalmoscopy and has five grades based on severity of disc swelling(4).

However, its reproducibility is limited, with inter-rater agreement as low as 49%, leading to significant variability in clinical practice (5).Clinically, a retinal photograph may not be obtained for most patients presenting to ophthalmology, whereas OCT is now ubiquitous in ophthalmological assessment and also commonly obtained at high street opticians. Artificial intelligence (AI) has been posed as a solution for consistent and scalable grading, with validation studies of bespoke deep learning models to identify papilloedema showing performance comparable to human experts (6–9).

Currently, most commercial OCT devices incorporate a high-contrast near-infrared reflectance (NIR) image acquired in combination with the structural OCT (10). NIR images are non-invasive and provide an *en face* composite image of multiple B scan OCTs which can be viewed as a fundus image and be used to grade papilloedema using the Frisen system (10). NIR images can also be easily obtained through an undilated pupil and can deliver adequate image quality even in poor signal quality and significant disc oedema (10–12).

The advent of automated machine learning (AutoML), also known as code-free AI, offers a promising solution to overcome the limitations of traditional grading systems. AutoML platforms enable the development of machine learning models for image classification without requiring expertise in coding, making them an attractive option for clinicians who are not versed in machine learning (ML) methods (13). Different AutoML platforms for classifying retinal photographs and OCT images have been shown to have varying performance and different features that increase usability, such as customising testing and training data splits (14).

This study is the first to demonstrate the feasibility of AutoML to develop a model to distinguish pseudopapilloedema from papilloedema using NIR images and compare the performance across different AutoML providers.

## Methods

### Dataset and Image Acquisition

Optic Nerve Head (ONH)-centred OCT and NIR images were acquired using the Heidelberg SPECTRALIS OCT imaging system (Heidelberg Engineering, Inc., Heidelberg, Germany) at the University Hospitals of Leicester NHS Trust between 2021 and 2024. Images were obtained under standardised acquisition protocols (acquisition software version 6.16.8.0), primarily employing two scanning approaches: (1) *peripapillary ring scans* with a circle diameter of 12° consisting of 768 A-scans per circle. Automatic real-time tracking (ART) mode was enabled, averaging 100 images to enhance image quality. These scans achieved axial scaling (Z-axis) of 3.87 µm/pixel and transverse scaling (X-axis) of approximately 15.13 µm/pixel and (2) *peripapillary volume scans* acquired under the high-speed mode, typically with a 15° × 15° volume pattern, comprising 37 horizontal B-scans. ART averaging of 54 images per B-scan was enabled. Enhanced depth imaging (EDI) was only used in suspected buried drusen cases. Near-infrared reflectance (NIR) images were obtained simultaneously with OCT scans with the following parameters: scan angle of 30°, image dimensions of 768 × 768 pixels (covering ∼8.1–10.9 mm², depending on axial length), transverse scaling of 10.61–14.15 µm/pixel, ART averaging of 45 frames per image, IR laser power at 100%, sensitivity (DC/DC) set between 81–90%, and auto-brightness enabled. All images were initially stored in the .E2E format, the proprietary file format used by the Heidelberg Engineering. To facilitate further analysis, these files were converted to fundus PNG images using the OCT-Convert Python package.

Patients were retrospectively identified using ICD-10 codes and electronic medical records. Diagnoses of papilloedema and ODD were clinically confirmed and verified through expert review of OCT B-scans and comprehensive medical record review. Longitudinal OCT and NIR images were included to represent varying Frisén grades, ensuring no duplication of identical grades for the same patient. Controls consisted of patients referred for ophthalmic evaluation unrelated to optic nerve pathology (e.g., posterior vitreous detachment, floaters etc). Detailed medical record review and OCT analysis confirmed no evidence of optic nerve pathology. For simplicity, this group is referred to as the “Normal” cohort throughout the study. The ground-truth dataset consisted of data from a total of 289 patients (813 images). 69 patients (185 images) were labelled normal, 85 patients (256 images) were labelled ODD, and 135 patients (372 images) were labelled papilloedema. (See supplementary Table 1 for demographic information.)

### Papilloedema grading

Images with papilloedema were graded using the Frisén system by three independent graders. A consensus grade was reached for each image, and severity was classified as mild/moderate (Frisén grades 1-3) or severe (Frisén grades 4-5). This grouped severity classification has been described in similar studies (9). The severity labels were used to train the AutoML models.

### Training and testing

Patients were randomly split into training (80%) and testing (20%) sets, ensuring that images from the same patient did not exist in different sets to develop two models. Testing and training data splits are shown in figure 1, where model 1 aimed to differentiate severity of papilloedema using the modified grading (Figure 1A) and model 2 aimed to differentiate papilloedema from ODD and normal optic discs (Figure 1B).

**Figure 1.**
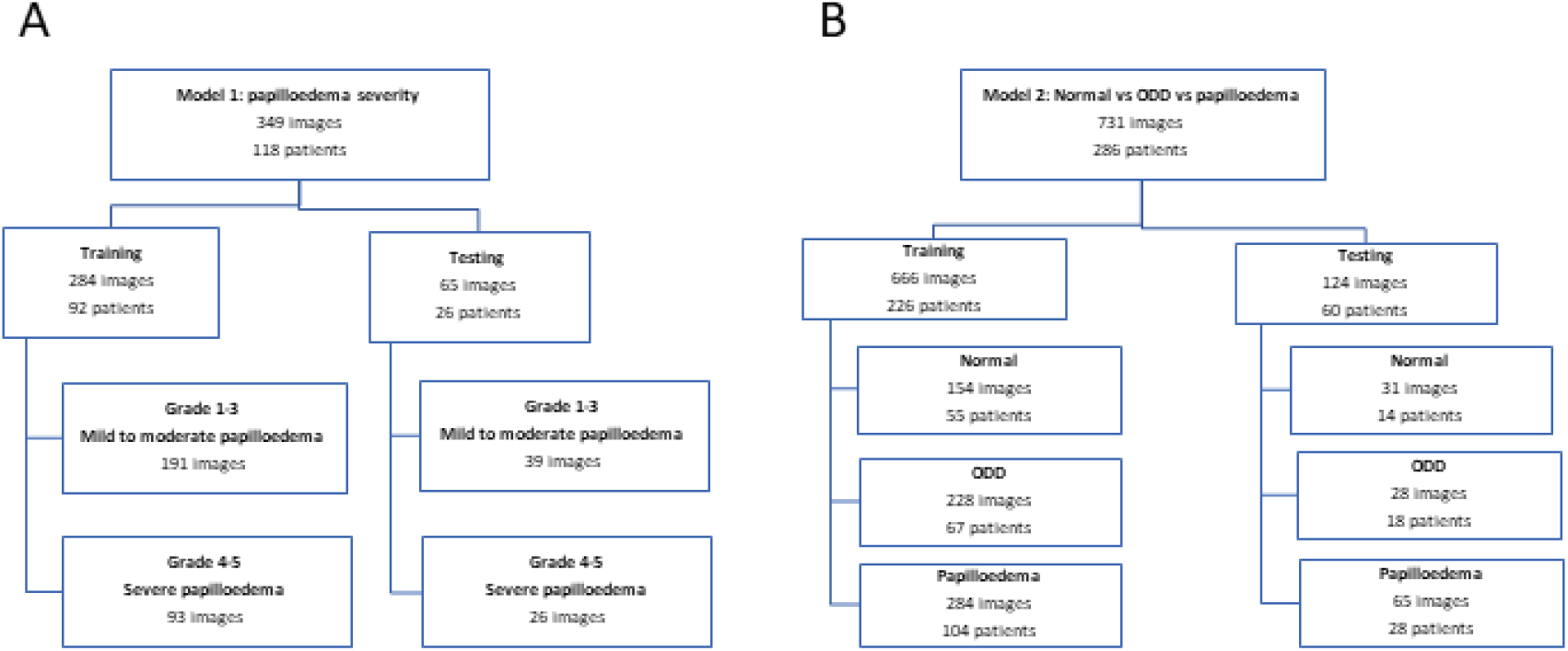
A) training and testing dataset split for Model 1: papilloedema severity mild/moderate vs severe and B) training and testing dataset split for Model 2, differentiating papilloedema vs normal disc vs ODD

### Code-free ML platform

The clinical research team with limited previous ML coding experience performed the AutoML model development. Amazon Rekognition (AWS), Medic Mind and Google Vertex platforms were selected for their ability to manually customise the training and testing data splits, which allowed data to be split at the patient level and prevented images from the same patient being used for testing and training the model. Furthermore, these platforms have previously been shown to perform well in image classification tasks using fundus photographs and OCT images (14). The models were evaluated using a custom script. These platforms only provide confidence score of each class and precision, recall, F1 score, and AUC are derived from this.

### Ethics approval

All data in this study was retrospectively collected, with necessary ethical approval (IRAS Project ID: 261121; REC Reference: 20/EM/0040)).

## Results

### Model 1: Papilloedema Severity Grading

All three models performed well in differentiating between mild to moderate and severe papilloedema. AWS model performed the best (AUC 0.94, F1 score 0.78), followed by Vertex (AUC 0.84, F1 score 0.74) and Medic Mind (AUC 0.82, F1 score 0.79) (Table 1). AWS most frequently misclassified mild to moderate papilloedema as severe papilloedema, however, Vertex and Medic Mind most frequently misclassified severe papilloedema as mild to moderate papilloedema (Figure 2).

**Figure 2:**
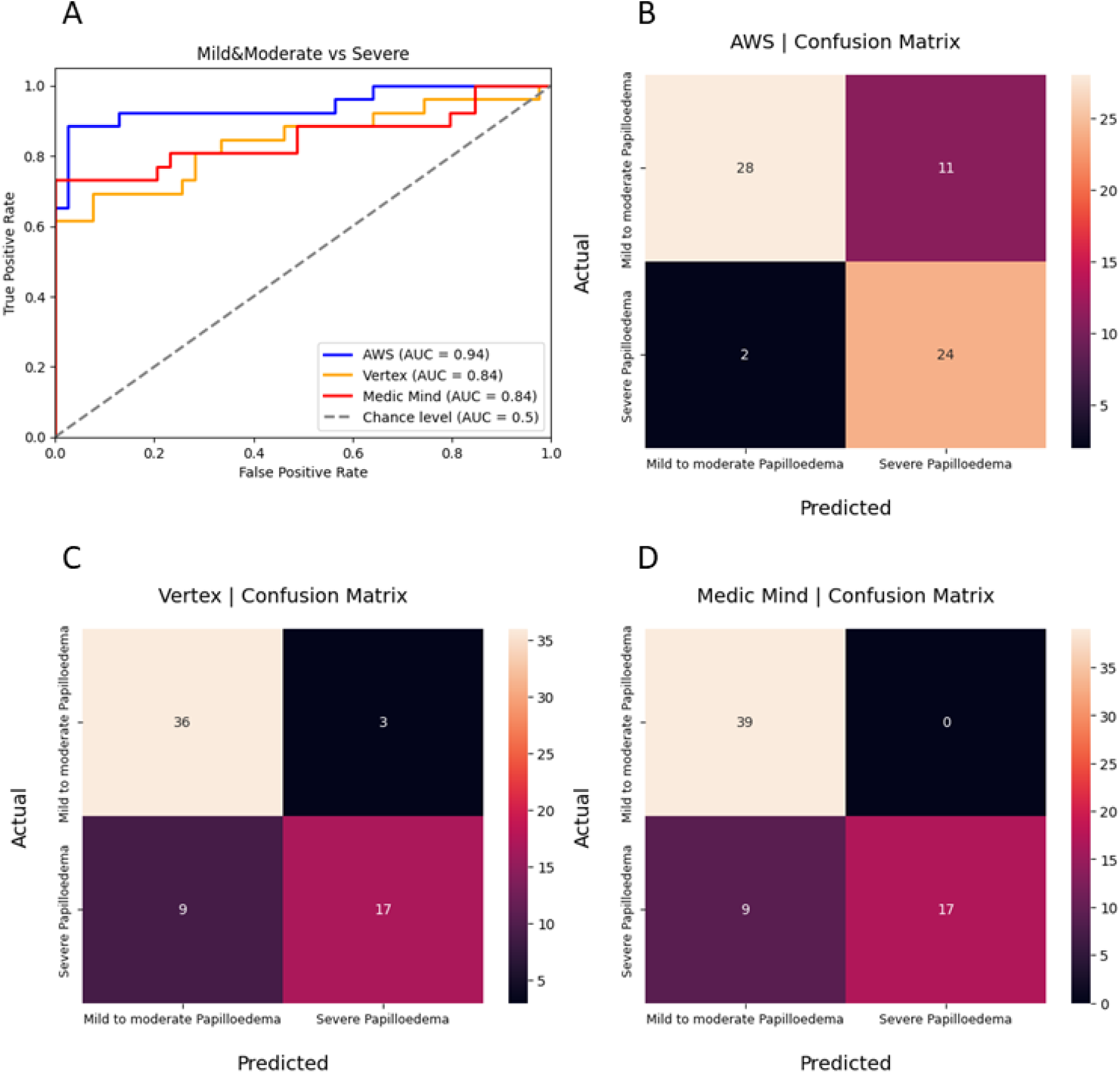
A) AUC curves of model 1 performance between Medic Mind, Google Vertex and AWS AutoML models in classifying papilloedema severity (mild/moderate vs severe), confusion matrices for model 1 on B) AWS, C) Vertex and D) Medic Mind

**Table 1:** Performance metrics including F1 score, precision and recall for model 1 (mild/moderate vs severe papilloedema) and model 2 (normal optic discs vs ODD vs papilloedema). ODD = Optic disc drusen.

|  | **F1** | **Precision** | **Recall** |
| --- | --- | --- | --- |
| *Model 1: Mild/moderate vs severe papilloedema* |  |  |  |
| AWS | 0.79 | 0.69 | 0.92 |
| Vertex | 0.74 | 0.85 | 0.65 |
| Medic Mind | 0.79 | 1.0 | 0.65 |
| *Model 2 Papilloedema vs ODD vs normal optic disc* |  |  |  |
| AWS | 0.81 | 0.81 | 0.84 |
| Vertex | 0.73 | 0.74 | 0.74 |
| Medic Mind | 0.72 | 0.74 | 0.78 |

### Model 2: Papilloedema vs ODD vs Normal

All three models demonstrated good performance in differentiating between normal optic discs, ODD and papilloedema in the multiclass model 2. AWS showed the best performance overall (average AUC 0.92, F1 score 0.81), closely followed by Vertex (average AUC 0.87, F1 score 0.73) and Medic Mind (average AUC 0.87, F1 score 0.72) (Table 1). Overall, the models from all three platforms showed the best performance in identifying normal optic discs (figure 3). Considering accurate identification of papilloedema would likely have the greatest clinical utility, AWS also showed the best performance in differentiating papilloedema (AUC 0.90), compared to Vertex (AUC 0.88) and Medic Mind (AUC 0.82) (Figure 3).

**Figure 3:**
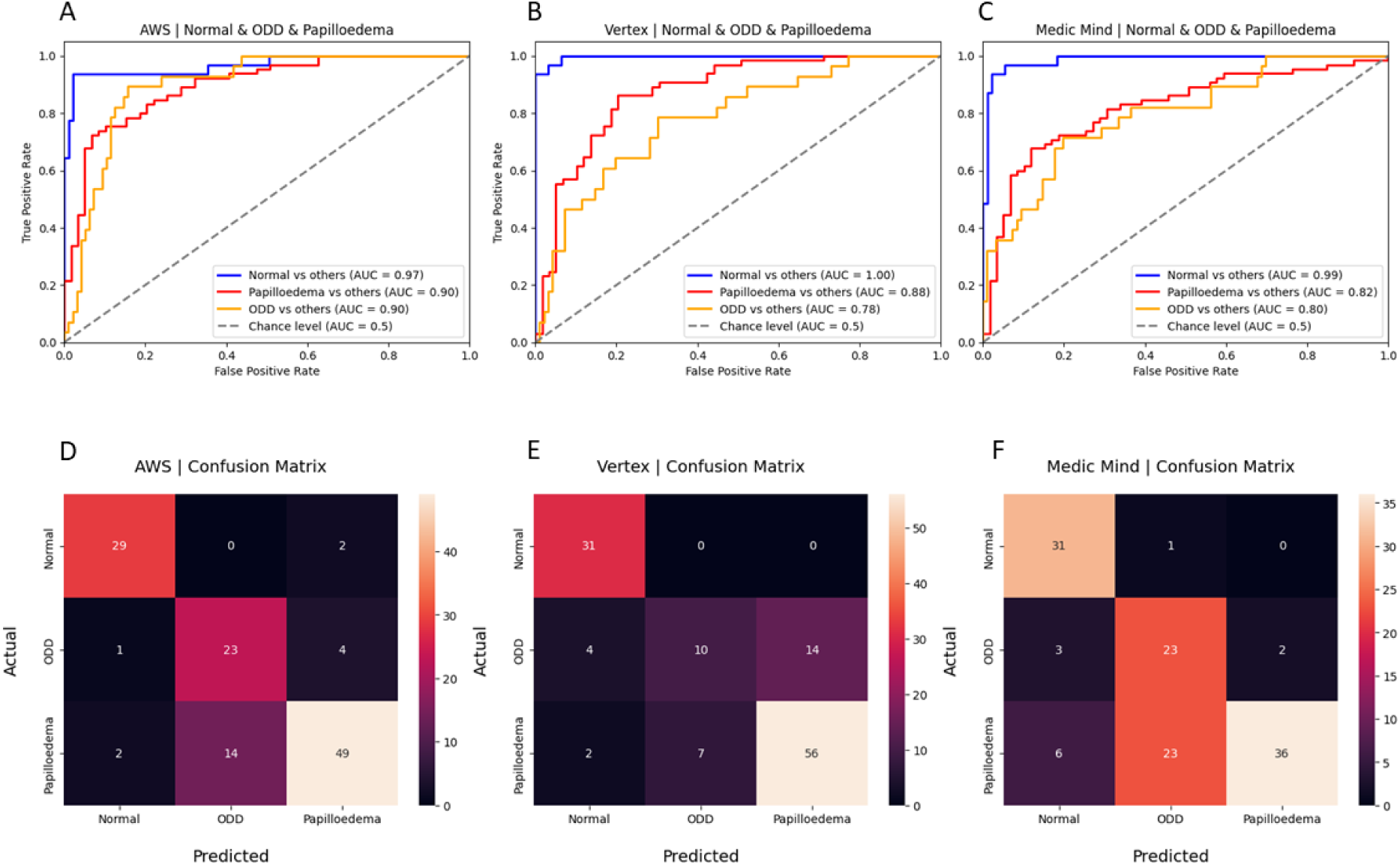
AUC curves of performance in classification model 2 (papilloedema vs normal optic discs vs ODD) for each platform A) AWS, B) Google Vertex and C) Medic Mind with corresponding confusion matrices for each model, D) AWS, E) Google Vertex and F) Medic Mind. ODD = Optic disc drusen, AWS = Amazon Rekognition

The models most frequently misclassified ODD, as papilloedema or normal optic discs (Figure 4a-c). Whilst both AWS and Vertex misclassified 2 out of 65 papilloedema images as normal optic discs, Medic Mind misclassified 6 papilloedema images as normal (Figure 3). Images of papilloedema misclassified as normal showed mild or moderate papilloedema (figure 4d-e) with one image showing an image of mild papilloedema in a patient who previously had severe papilloedema (figure 4f).

**Figure 4:**
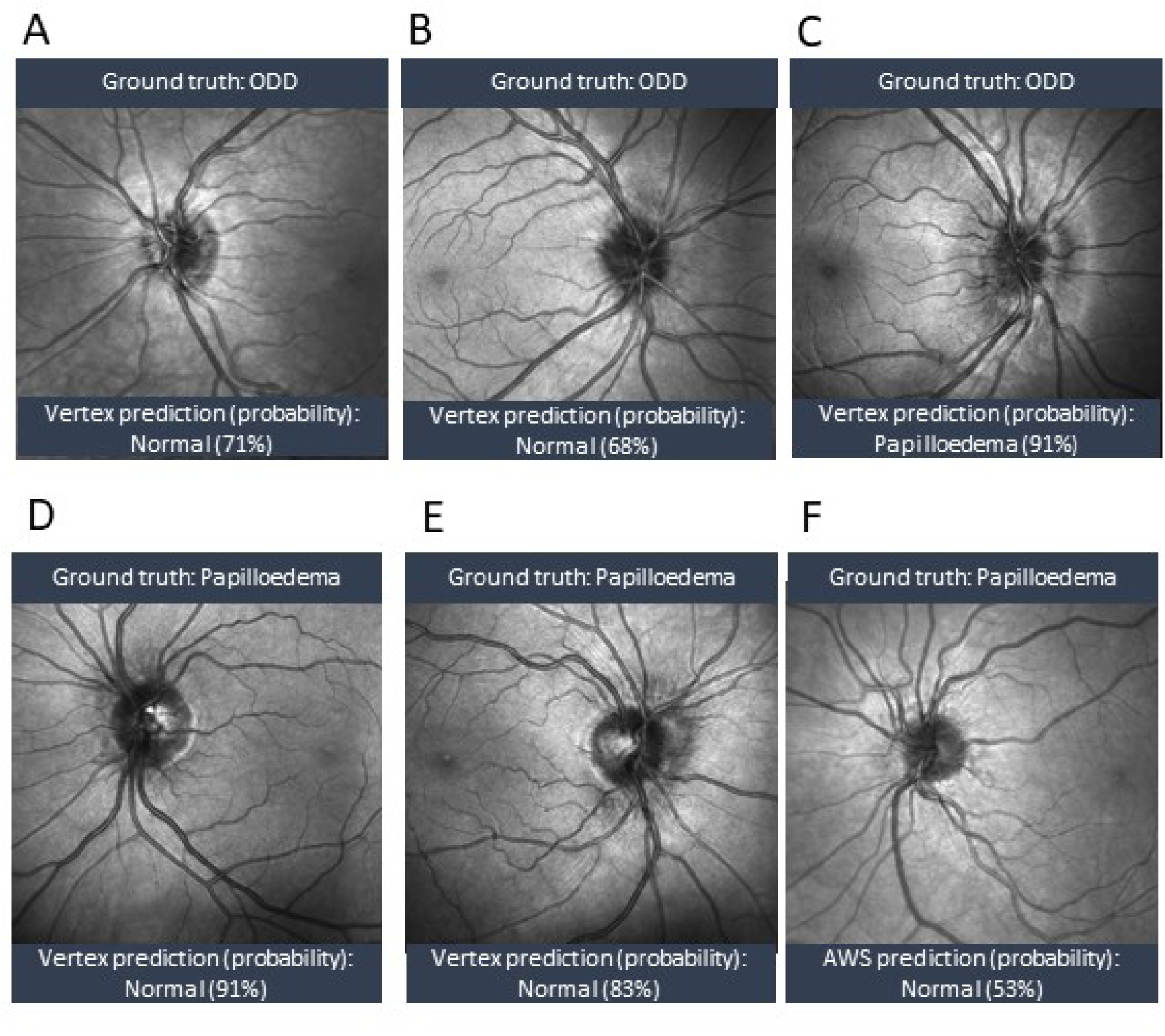
A-F examples of misclassified images with ground truth label and predicted label and model probability of prediction. ODD = Optic disc drusen, AWS = Amazon Rekognition

## Discussion

This study is the first to demonstrate the feasibility of using AutoML platforms to classify and grade papilloedema from NIR OCT images. In this study, two models have successfully been developed using three different AutoML platforms - AWS, Google Vertex and Medic Mind - to improve recognition and characterisation of papilloedema using NIR images. Across all three platforms, the binary classification model was reliably able to predict severity of papilloedema and the multiclass model showed good reliability in differentiating papilloedema from normal optic discs, but performed least reliably in predicting ODD from papilloedema with higher rates of misclassification.

To our knowledge, there is no published comparison of AutoML models for classifying papilloedema based on NIR images. Whilst previous studies have shown custom engineered deep learning systems (DLS) can achieve high accuracy (AUC 0.89–0.99) in detecting papilloedema from fundus images, developing DLS requires significant computational resources, technical expertise, and extensive annotated datasets, which limit their widespread implementation (15–19).

Difficulty in classifying papilloedema using Frisén grading has been noted, particularly surrounding the lack of reproducibility and utility in monitoring optic disc changes (20, 21). Indeed, in clinical practice, the simplified classification of papilloedema used in this study may often be used in clinical decision making (9). Fundus findings on examination may also be triangulated with patient symptoms and other imaging modalities, including quantitative and qualitative findings on transverse axial OCT images, and ultrasonography findings (22).

While AutoML models have been shown to be comparable to bespoke DLS in ophthalmology, it is unclear how the inherent challenges of using the Frisén grading scale are reflected in the AutoML model performance, which is reliant on labelled data by human graders (23). When training the BONSAI model to determine severity of papilloedema, moderate papilloedema (Frisén grade 3) was most frequently misclassified as severe papilloedema, and misclassified cases of severe papilloedema contained other optic disc pathologies such as cotton wool spots or haemorrhages (9).

In our study, the AutoML models performed well with high AUCs for classifying mild to moderate vs severe papilloedema, though there was variability in misclassifications between platforms.

Image type and quality may impact model performance, though the BONSAI DLS has been shown to have comparable performance in both dilated fundus photographs and undilated fundus photographs, obtained by emergency department professionals, with 84% sensitivity and 99% specificity for differentiating papilloedema from normal optic discs and ODD (16, 17). To ensure sufficient image quality, further DLS models have been proposed to classify image quality of optic discs in fundus photographs (24).

As part of data curation for model development, images with decentration or poor quality are excluded as ungradable. The original validation study of the BONSAI DLS excluded 153 photographs deemed ungradable, with 15,846 images subsequently included (16). In comparison, a much higher rate of ungradable images was reported in the study using undilated fundus photographs obtained by nurses or students in the emergency department where 463 patients’ images were excluded from the original 1291 patients due to poor quality image or missing the optic disc (17).

Conversely, the low rate of ungradable images in our study is notable, attesting to the reliable quality NIR image generated by commercial OCT. These findings could support the use of NIR images to assess the optic disc in low-resource settings, where one machine could reliably produce images of sufficient quality for both transverse axial and *en face* images of the retina. Moreover, the use of NIR images provides a compelling case for their utility in both high-resource and low-resource settings. These images, with no flash or pupil dilation requirements, are particularly advantageous for paediatric patients or individuals with photophobia, where traditional fundus photography may fail due to artefacts like blink shadows or iris silhouettes.

Strengths of this study include manual data splitting at a patient level, using longitudinal data and having a multi-rater consensus to establish ground truth labels. Ensuring images from the same patient did not appear in both training and testing sets reduced the risk of overestimation of the model performance and prevented inadvertent data leakage between sets. Using longitudinal data, ensuring only different Frisén graded images were used for any single patient, enhanced the sample size without duplicating images of the same grade, which could otherwise lead to model overfitting.

Limitations of the dataset include a relatively small sample size, class imbalance and lack of external validation. The sample may not have captured the breadth of phenotypes of optic discs, ODD and papilloedema. Furthermore, the ODD class had the lowest sample size which could have contributed to the model’s performance for this class. The size of the dataset may have also contributed to model overfitting which would need to be evaluated by testing on a new, unseen dataset. External validation was not carried out on this model, which would be necessary to assess the utility of the models in different clinical settings.

This study highlights the feasibility and potential of AutoML platforms in grading papilloedema severity and classifying NIR images into normal, ODD, and papilloedema categories. By providing accessible tools for AI model development, AutoML has the potential to transform the diagnostic landscape of neuro-ophthalmology, particularly in settings with limited resources, combined with clinician input to detect misclassified true papilloedema. Future research should focus on addressing dataset limitations and externally validating the models to assess generalisability and maximise clinical impact.

## Data Availability

All data produced in the present study are available upon reasonable request to the authors

---

**Supplementary table 1:** demographic information for patients included in the ground-truth dataset.

|  | **Papilloedema** | **ODD** | **Controls** |
| --- | --- | --- | --- |
| **Total Participants** | 135 | 85 | 69 |
| **Age (Mean ± SD)** | 30.2 ± 12.4 | 25.3 ± 14.7 | 49.4 ± 19.0 |
| **M:F Ratio** | 23:112 | 33:52 | 31:38 |
| **Ethnicity Distribution** | White: 106,<br>Asian: 18,<br>Mixed: 6,<br>Other: 3,<br>Black: 2 | White: 78,<br>Asian: 6,<br>Mixed: 1 | White: 35,<br>Asian: 16,<br>Black: 14,<br>Mixed: 3,<br>Other: 1 |

